# Immunogenicity of the ChAdOx1 nCoV-19 and the BBV152 Vaccines in Patients with Autoimmune Rheumatic Diseases

**DOI:** 10.1101/2021.06.06.21258417

**Authors:** Padmanabha Shenoy, Sakir Ahmed, Somy Cherian, Aby Paul, Veena Shenoy, Anuroopa Vijayan, Reshma Reji, Arya Thampi, AS Sageer Babu, Manju Mohan

**Author notes:** Corresponding Author : Dr Padmanabha Shenoy, Centre for arthritis and rheumatism excellence (CARE), Nettoor, Kochi, Kerala 682040, India.

## Abstract

**Introduction:** There is limited information on the effectiveness of COVID-19 vaccination in patients with autoimmune rheumatic diseases (AIRD).

**Methods:** 136 consecutive patients with rheumatic diseases who never had a diagnosis of COVID-19 previously, and had completed vaccination with either the ChAdOx1 or BBV152 vaccines were recruited. Their IgG antibody titres to the Spike protein were estimated 1 month after the second dose.

**Results:** 102 patients had AIRD while the 34 had non-AIRD. Lesser patients with AIRD (92/102) had positive antibodies titres than ones with non-AIRD(33/34) [p<0.001]. Amongst patients who received the ChAdOX1 vaccine, the AIRD group had lower antibody titres. Although the AIRD patients receiving BBV152 had similarly lower titres numerically, this did not attain statistical significance probably due to lesser numbers. Comparing the two vaccines, 114(95%) of those who received ChAdOx1 (n=120) and 11(68.7%) of those who received BBV152(n=16) had detectable antibodies [p=0.004]. Antibody titres also were higher in ChAdOx1 recipients when compared to BBV152.

To validate the findings, we estimated antibody titres in 30 healthy people each who had received either vaccine. All 30 who had received ChAdOX1 and only 23/30 of those who had received BBV152 had positive antibodies (p=0.011).

**Conclusion:** In this preliminary analysis, patients with AIRD had lower seroconversion rates as well as lower antibody titres as compared to patients with non-AIRD. Also,the humoral immunogenicity of the BBV152 vaccine appears to be less than that of the ChAdOX1 vaccine. Validation using larger numbers and testing of cellular immunity is urgently required.

---

There is limited information on the effectiveness of COVID-19 vaccination in patients with autoimmune rheumatic diseases (AIRD). Some data is emerging for the mRNA vaccines[1,2], but less data is available for the non-replicating adenoviral vector ChAdOx1 nCoV-19 vaccine and the Indian heat-killed BBV152 vaccine. We have collated data that both vaccines were safe in 725 patients with rheumatic and musculoskeletal diseases(RMDs).[3]

With ethics clearance (Institutional Ethics Committee of Sree Sudheendra Medical Mission Hospital No: IEC/2021/35) and written informed consent, we recruited a convenient sample of 136 RMD patients who never had a diagnosis of COVID-19 previously, and had completed two doses of either the ChAdOx1 or BBV152 vaccines. Around one month after the second dose, the presence of IgG antibody directed against the S1 domain of the SARS-CoV-2 spike protein was estimated using a commercial Chemiluminescent assay (Roche, Switzerland). The normality of data was checked by the Shapiro-Wilk test. The difference in proportions was tested by the Fisher Exact test and the difference in antibody titres was tested by Independent sample t-test after log transformation. P<0.05 was the cut-off for significance.

Out of the 136 patients, 102 had AIRD while the 34 others had non-AIRD. A significantly [p<0.001] lower proportion of patients with AIRD (92/102) had positive antibodies titres than ones with non-AIRD(33/34) [Supplementary Table 1]. The characteristics of patients who had a negative response to either vaccine are summarized in Supplementary tables 2. Amongst patients who received the ChAdOX1 vaccine, the AIRD group had lower antibody titres than the others. Although the AIRD patients receiving BBV152 had similarly lower numerical titres, this did not attain statistical significance probably due to lesser numbers [Supplementary table 1]

Comparing the two vaccines, 114(95%) of those who received ChAdOx1 (n=120) and 11(68.7%) of those who received BBV152(n=16) had detectable antibodies [p=0.004]. Antibody titres also were higher in ChAdOx1 recipients when compared to BBV152. [Figure 1].

**Figure 1:**
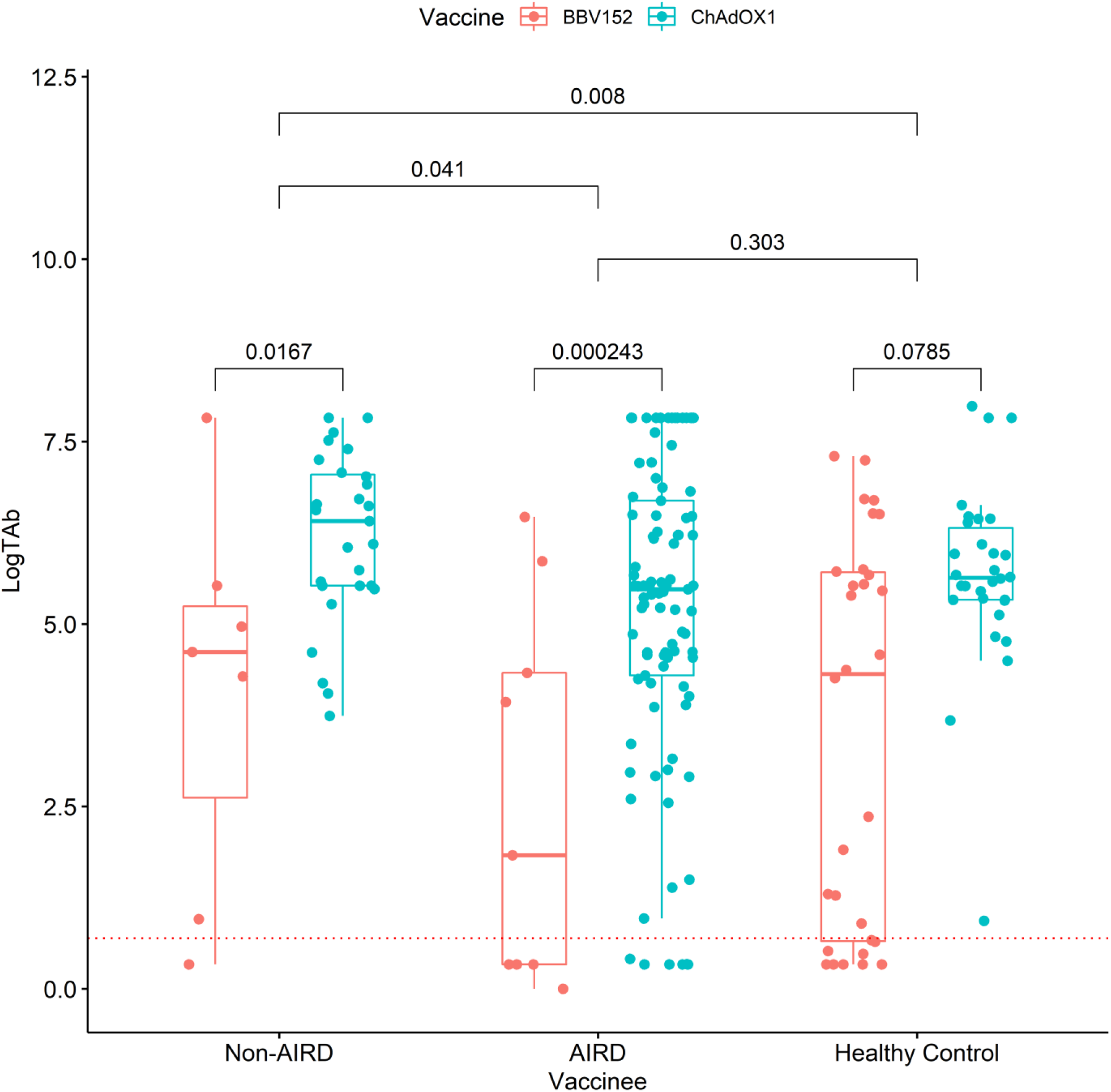
Log transformed antibody titres post vaccination with BBV152 versus ChAdOX1 vaccines, stratified according to: patients with non-autoimmune rheumatic diseases (Non-AIRD), patients with autoimmune rheumatic diseases (AIRD) and healthy controls. The three reference bars at the top show statistical difference between these three groups combing titres of both vaccines.

Thus, this preliminary analysis showed that patients with AIRD had lesser antibody positivity with lower titres than patients with non-AIRD. Also, the use of the BBV152 vaccine was associated with lower seroconversion rates and lower antibody titres as compared to the ChAdOX1 vaccine. To validate the findings, we estimated antibody titres in 30 healthy people each who had received either vaccine. All 30 who had received ChAdOX1 and only 23/30 of those who had received BBV152 had positive antibodies (p=0.011)[Supplementary Table 3]. Moreover, with in one month of antibody levels estimation, 3 out of the 7 controls who had negative antibodies had symptomatic COVID-19 infection.

Previously, seroconversion rates with the BNT162b2 mRNA vaccine have been shown to be lower in patients with immune-mediated inflammatory diseases [2]. A case series of RMD patients who had not developed antibodies post-inoculation with the mRNA vaccines, has shown most of these patients of lupus, myositis or vasculitis and were on rituximab or mycophenolate [4]. In our cohort, findings are similar for patients who had received ChAdOx1, but not BBV152. Patients with solid organ transplants have 30-50% seropositivity after mRNA vaccines[5,6]. Factors associated with poor seroconversion included age, steroid dose, triple immunosuppression and mycophenolate use [6]. Thus, specialized strategies are required for immunocompromised patients.

The limitations of this study include a non-random sample, a small sample size, lack of T cell immunogenicity data and that the pre-vaccination antibody status of the patients was not known. However, all these patients had denied symptomatic infection or test positivity.

Thus, seroconversion rates were lower in patients with AIRD than in those with non-AIRD. The humoral immunogenicity of the BBV152 vaccine appears to be less than that of the ChAdOX1 vaccine, especially in patients with AIRD. Validation using larger numbers, testing of cellular immunity and analysis of the effects of underlying disease and immunosuppressants is required. These findings may have practical implications in vaccination recommendations for patients with rheumatic diseases.

## Data Availability

All data is available and all authors have seen it

---

**Supplementary Table 1:** Characteristics of the cohort

| Parameters |  | AIRD(N=102) | Non AIRD(N=34) | Test for significance<br>(P value) |
| --- | --- | --- | --- | --- |
| Age (Mean(SD)) |  | 52(12.33) | 54.12(11.21) | 0.377 |
| Female |  | 81(79.40%) | 24(70.60%) | 0.241 |
| Comorbidities (at least one) |  | 37(36.30%) | 14(41.20%) | 0.684 |
| Vaccine received | ChAdOx1 | 93(91.20%) | 27(79.40%) | 0.120 |
|  | BBV152 | 9(8.8%) | 7(20.58%) |  |
| Time interval between 2<sup>nd</sup> dose and blood sampling(Mean(SD)) |  | 27.60(11.67) | 27.52(9.49) | 0.566 |
| Positive antibody titres post-vaccination |  | 92(90.2%) | 33(97.0%) | <0.001** |
| Anti-SARS CoV-2 Antibody levels<br>(Median (IQR)) |  | 223.60(53.06-656.40) | 366.25(131.50-1134.50) | 0.303 |
| Anti-SARS CoV-2 Antibody levels stratified by vaccine<br>(Median (IQR)) | ChAdOx1 | 238(70.46-825.5) | 608(250-1178) | 0.032* |
|  | BBV152 | 5.24(0.4-212.5) | 100(1.6-250) | 0.241 |

**Supplementary table 2:** List of AIRD patients who did not have a response to either vaccine.

| Cas<br>e | Ag<br>e | Gende<br>r | Diagnosis | Co-Morbidity | Vaccine | Time<br>From<br>dose 2<br>to<br>Antibod<br>y<br>testing | DMARDS<br>/Immuno-<br>suppressants /<br>Biologics | Daily steroid<br>dose in<br>prednisolone<br>equivalent(m<br>g) |
| --- | --- | --- | --- | --- | --- | --- | --- | --- |
| 1 | 67 | Femal<br>e | Rheumatoid<br>Arthritis | Diabetes<br>mellitus type<br>2 | ChAdOx<br>1 | 18 | Methotrexate,<br>Leflunomide,<br>Rituximab | 2.5 |
| 2 | 57 | Femal<br>e | Rheumatoid<br>Arthritis | Hypertension | ChAdOx<br>1 | 29 | Hydroxychloroqui<br>ne<br>Iguratimod,<br>Rituximab | 0 |
| 3 | 68 | Femal<br>e | Systemic<br>Lupus<br>Erythematos<br>is | Diabetes<br>mellitus type<br>2 | ChAdOx<br>1 | 30 | MMF, Rituximab | 0 |
| 4 | 40 | Femal<br>e | Systemic<br>sclerosis | Nil | ChAdOx<br>1 | 53 | MMF | 0 |
| 5 | 51 | Femal<br>e | Dermato-<br>myositis | Hypothyroidis<br>m | ChAdOx<br>1 | 22 | MMF | 25 |
| 6 | 52 | Femal<br>e | Rheumatoid<br>Arthritis | Diabetes<br>mellitus type<br>2 | ChAdOx<br>1 | 62 | Methotrexate,<br>Hydroxychloroqui<br>ne | 0 |
| 7 | 40 | Male | Palindromic<br>Rheumatism | Nil | BBV152 | 24 | Adalimumab,<br>Hydroxychloroqui<br>ne | 0 |
| 8 | 36 | Femal<br>e | Polyarteritis<br>Nodosa | Nil | BBV152 | 44 | Methotrexate | 5 |
| 9 | 55 | Femal<br>e | Rheumatoid<br>Vasculitis | Diabetes<br>mellitus type<br>2 | BBV152 | 49 | MMF,<br>Hydroxychloroqui<br>ne | 0 |
| 10 | 42 | Femal<br>e | Systemic<br>sclerosis | Dyslipidaemia | BBV152 | 21 | Tacrolimus | 5 |

MMF: Mycophenolate

**Supplementary Table 3:** Characteristics of the Healthy Controls

| Parameters | ChAdOx1<br>N=30 | BBV152<br>N=30 | P<br>value |
| --- | --- | --- | --- |
| Age (Mean(SD)) | 43.60(9.17) | 44.20(7.48) | 0.782 |
| Male | 28(93.33%) | 28(93.33%) | 1 |
| Comorbidities (at least one) | 12(40%) | 10(33.33%) | 0.789 |
| Time interval between 2<sup>nd</sup> dose and blood sampling<br>(Mean(SD)) | 43(10.60) | 43.33(7.26) | 0.888 |
| Positive antibody titres post-vaccination | 30(100%) | 23(76.66%) | 0.011 |
| Anti-SARS CoV-2 Antibody levels<br>(Median (IQR)) | 278(205-603.12) | 73.89(0.85-306.25) | 0.07 |

**Supplementary Table 4:**
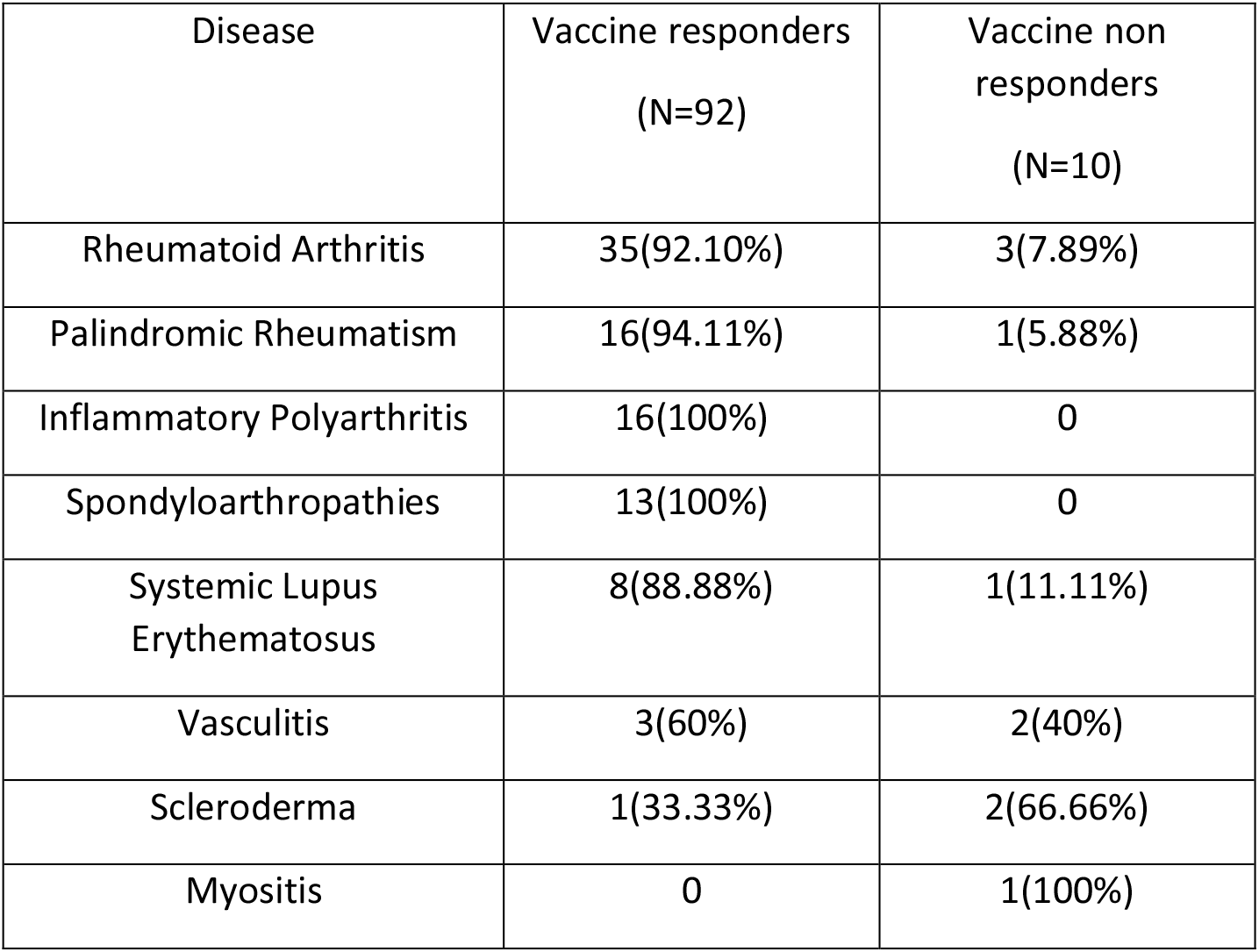
Distribution of different autoimmune rheumatic diseases amongst vaccine responders versus non-responders

| Disease | Vaccine responders<br>(N=92) | Vaccine non<br>responders<br>(N=10) |
| --- | --- | --- |
| Rheumatoid Arthritis | 35(92.10%) | 3(7.89%) |
| Palindromic Rheumatism | 16(94.11%) | 1(5.88%) |
| Inflammatory Polyarthrititis | 16(100%) | 0 |
| Spondyloarthropathies | 13(100%) | 0 |
| Systemic Lupus<br>Erythematosus | 8(88.88%) | 1(11.11%) |
| Vasculitis | 3(60%) | 2(40%) |
| Scleroderma | 1(33.33%) | 2(66.66%) |
| Myositis | 0 | 1(100%) |

**Supplementary Table 5:**
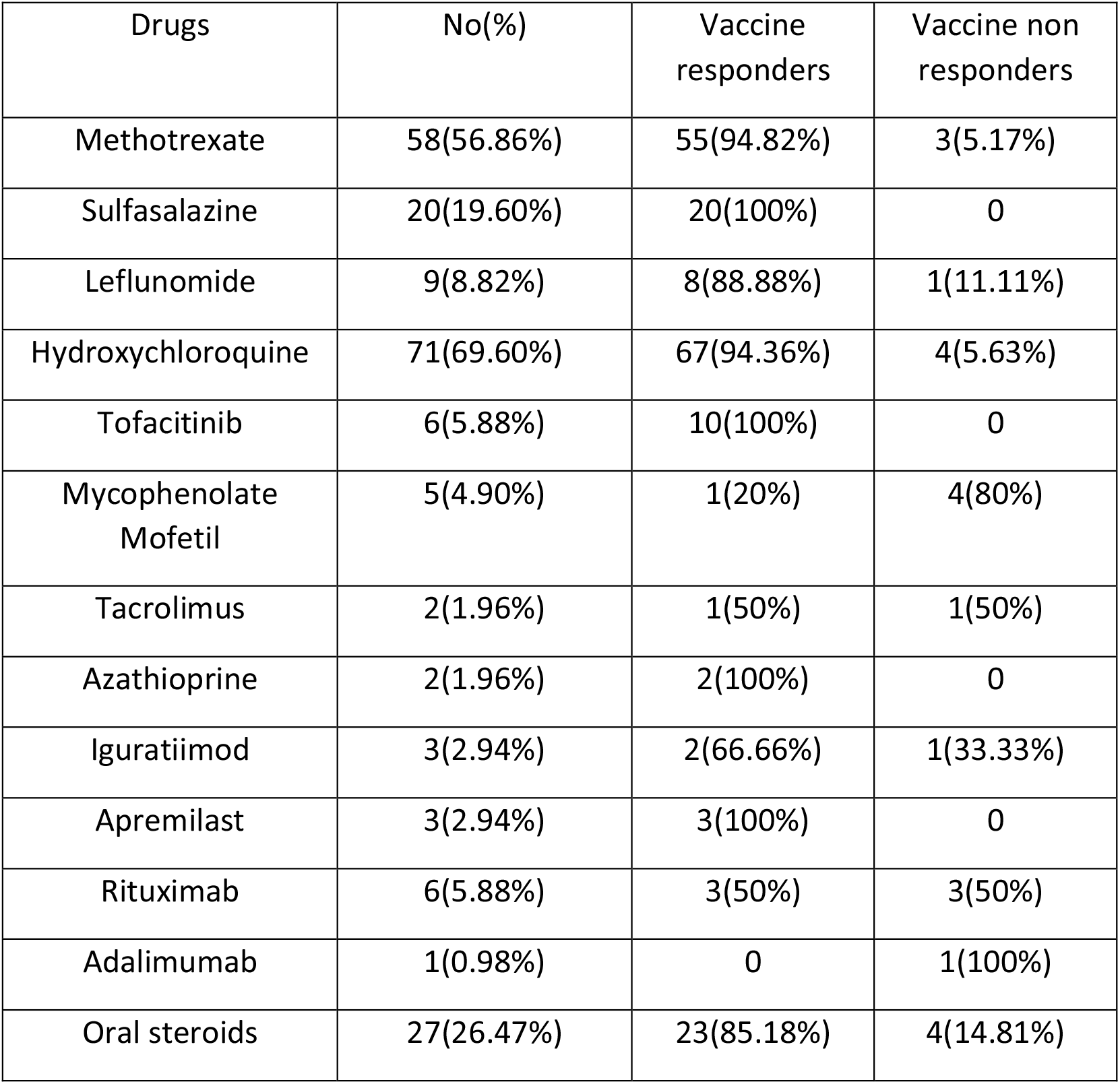
Use of immunosuppressant drugs amongst vaccine responders versus non-responders

## References

1 Geisen UM, Berner DK, Tran F, et al. Immunogenicity and safety of anti-SARS-CoV-2 mRNA vaccines in patients with chronic inflammatory conditions and immunosuppressive therapy in a monocentric cohort. Annals of the Rheumatic Diseases Published Online First: 24 March 2021. doi:10.1136/annrheumdis-2021-220272

2 Simon D, Tascilar K, Fagni F, et al. SARS-CoV-2 vaccination responses in untreated, conventionally treated and anticytokine-treated patients with immune-mediated inflammatory diseases. Ann Rheum Dis Published Online First: 6 May 2021. doi:10.1136/annrheumdis-2021-220461

3 Cheriyan S, Paul A, Ahmed S, et al. Safety of the ChAdOx1 nCoV-19 and the BBV152 Vaccines in 724 Patients having Rheumatic Diseases. Rochester, NY: : Social Science Research Network 2021. https://papers.ssrn.com/abstract=3853696 (accessed 28 May 2021). [Article accepted by Rheumatol Int: we will update the reference once the peer-reviewed version is online].

4 Connolly CM, Boyarsky BJ, Ruddy JA, et al. Absence of Humoral Response After Two-Dose SARS-CoV-2 Messenger RNA Vaccination in Patients With Rheumatic and Musculoskeletal Diseases: A Case Series. Ann Intern Med Published Online First: 25 May 2021. doi:10.7326/M21-1451

5 Marion O, Del Bello A, Abravanel F, et al. Safety and Immunogenicity of Anti–SARS-CoV-2 Messenger RNA Vaccines in Recipients of Solid Organ Transplants. Ann Intern Med Published Online First: 25 May 2021. doi:10.7326/M21-1341

6 Grupper A, Rabinowich L, Schwartz D, et al. Reduced humoral response to mRNA SARS-CoV-2 BNT162b2 vaccine in kidney transplant recipients without prior exposure to the virus. Am J Transplant Published Online First: 18 April 2021. doi:10.1111/ajt.16615

